# Multiomic Analyses Direct Hypotheses for Creutzfeldt-Jakob Disease Risk Genes

**DOI:** 10.1101/2024.07.19.24310476

**Authors:** Fahri Küçükali, Elizabeth Hill, Tijs Watzeels, Holger Hummerich, Tracy Campbell, Lee Darwent, Steven Collins, Christiane Stehmann, Gabor G Kovacs, Michael D Geschwind, Karl Frontzek, Herbert Budka, Ellen Gelpi, Adriano Aguzzi, Sven J van der Lee, Cornelia M van Duijn, Pawel P Liberski, Miguel Calero, Pascual Sanchez-Juan, Elodie Bouaziz-Amar, Jean-Louis Laplanche, Stéphane Haïk, Jean-Phillipe Brandel, Angela Mammana, Sabina Capellari, Anna Poleggi, Anna Ladogana, Maurizio Pocchiari, Saima Zafar, Stephanie Booth, Gerard H Jansen, Aušrinė Areškevičiūtė, Eva Løbner Lund, Katie Glisic, Piero Parchi, Peter Hermann, Inga Zerr, Jiri Safar, Pierluigi Gambetti, Brian S Appleby, John Collinge, Kristel Sleegers, Simon Mead

## Abstract

Prions are assemblies of misfolded prion protein that cause several fatal and transmissible neurodegenerative diseases, with the most common phenotype in humans being sporadic Creutzfeldt-Jakob disease (sCJD). Aside from variation of the prion protein itself, molecular risk factors are not well understood. Prion and prion-like mechanisms are thought to underpin common neurodegenerative disorders meaning that the elucidation of mechanisms could have broad relevance. Herein we sought to further develop our understanding of the factors that confer risk of sCJD using a systematic gene prioritization and functional interpretation pipeline based on multiomic integrative analyses. We integrated the published sCJD genome-wide association study (GWAS) summary statistics with publicly available bulk brain and brain cell type gene and protein expression datasets. We performed multiple transcriptome and proteome-wide association studies (TWAS & PWAS) and Bayesian genetic colocalization analyses between sCJD risk association signals and multiple brain molecular quantitative trait loci signals. We then applied our systematic gene prioritization pipeline on the obtained results and nominated prioritized sCJD risk genes with risk-associated molecular mechanisms in a transcriptome and proteome-wide manner. Genetic upregulation of both gene and protein expression of syntaxin-6 (*STX6*) in the brain was associated with sCJD risk in multiple datasets, with a risk-associated gene expression regulation specific to oligodendrocytes. Similarly, increased gene and protein expression of protein disulfide isomerase family A member 4 (*PDIA4*), involved in the unfolded protein response, was linked to increased disease risk, particularly in excitatory neurons. Protein expression of mesencephalic astrocyte derived neurotrophic factor (*MANF*), involved in protection against endoplasmic reticulum stress and sulfatide binding (linking to the enzyme in the final step of sulfatide synthesis, encoded by sCJD risk gene *GAL3ST1*), was identified as protective against sCJD. In total 32 genes were prioritized into two tiers based on level of evidence and confidence for further studies. This study provides insights into the genetically-associated molecular mechanisms underlying sCJD susceptibility and prioritizes several specific hypotheses for exploration beyond the prion protein itself and beyond the previously highlighted sCJD risk loci through the newly prioritized sCJD risk genes and mechanisms. These findings highlight the importance of glial cells, sulfatides and the excitatory neuron unfolded protein response in sCJD pathogenesis.

## Introduction

Prions are infectious, proteinaceous pathogens composed of fibrillar assemblies of misfolded forms of host-encoded prion protein (PrP)^1^. Prions replicate by templated misfolding leading to fibril growth and fission^2^. Prion propagation leads to the generation of neurotoxic species and neurodegeneration. This underlying molecular mechanism is at the core of a multitude of human and animal prion diseases, and several aspects of the mechanism (so-called “prion-like”) are shared with the more common neurodegenerative disorders^2^.

Human prion diseases are unusual amongst neurodegenerative diseases in having three different types of aetiology: as well as arising due to rare pathogenic mutations in *PRNP* encoding PrP^C^ (inherited prion disease accounting for ∼10-15% cases) and spontaneously (sporadic prion disease accounting for ∼85% cases), the disease can also be acquired through transmission between humans or zoonotically(<1% cases)^3–5^. Sporadic Creutzfeldt-Jakob disease (sCJD) is the most common human prion disease, which has a lifetime risk of ∼1:5000^6^, and typically presents as a rapidly progressing dementia. There are no established disease-modifying treatments for sCJD although treatments targeting PrP using different therapeutic modalities such as employing PrP-targeting monoclonal antibodies have been reported ^7^ and *PRNP*-targeting ASOs (Phase 1/2a trial employing ION717, NCT06153966) are under investigation. Currently however the diseases are universally fatal and, for optimal disease mitigation, new therapeutic targets may be required beyond PrP itself.

In 2020, a collaborative genome-wide association study (GWAS) was conducted in sCJD, which identified novel risk loci for sCJD susceptibility^8^. In addition to the well-known variants in the *PRNP* gene, this study independently replicated findings at two further novel loci, at or within *STX6* and *GAL3ST1,* to be associated with sCJD risk. *STX6* encodes syntaxin-6, a SNARE protein predominantly involved in retrograde trafficking from early endosomes to the *trans-*Golgi network^9,10^, implicating intracellular trafficking as a causal molecular pathway in sCJD. *GAL3ST1* encodes galactose-3-O-sulfotransferase 1 predominantly in oligodendrocytes, the exclusive enzyme involved in the final step of sulfatide synthesis, which is a key constituent of the myelin sheath^11^. Two other genes were implicated in sCJD risk by tests that summarise evidence for association across the entire gene locus, including *PDIA4*, and variants in and near to a further gene, *BMERB1*, which were very close to genome-wide thresholds of association^8^.

We aimed to harness transcriptomic and proteomic datasets to provide further insight into sCJD risk in studies such as transcriptome-wide association studies (TWAS) and proteome-wide association studies (PWAS), respectively when integrated with the genetic datasets. Herein, the latest sCJD GWAS summary statistics^8^ were integrated with functional annotations (expression quantitative trait loci [eQTL] and protein expression QTL [pQTL]) to infer genetic up- and down-regulation of genes and/or protein expression in brain regions and associated with sCJD susceptibility. As the approach in TWAS/PWAS combines associations across variants, thus reducing the multiple testing burden, these analyses offer a powerful, complementary approach to conventional GWAS to develop supporting or negating evidence for loci that were subthreshold (*PDIA4, BMERB1)* or loci that did not reach the genome-wide significant threshold in the previous GWAS^8^. Furthermore, it allows exploration of expression-related genetic mechanisms underlying the GWAS association signals already identified (*PRNP, STX6, GAL3ST1)* uncovering further mechanistic insights into sCJD risk loci, in addition to nominating new TWAS/PWAS significant prioritized risk genes within subthreshold loci for generating novel disease-relevant hypotheses. Importantly, there are precedents of similarly designed studies achieving these goals in other neurological diseases^12–18^.

This work provides compelling evidence for risk variants in and around the *STX6* locus driving increased transcript and protein expression in the brain and consequently disease risk, which intriguingly and unexpectedly predominates in oligodendrocytes. This study also prioritizes the previous subthreshold GWAS hit, *PDIA4,* which is involved in the unfolded protein response (UPR), as being implicated in sCJD susceptibility, driven by *PDIA4* upregulation. Interestingly, this effect seemed to localise to excitatory neurons with interactions with the PWAS hit, *MANF*, providing an intriguing link to sulfatide metabolism and *GAL3ST1*. Several other subthreshold hits were also identified with potential relevance to prion disease mechanisms, including the previously identified subthreshold GWAS hit, *BMERB1*.

Taken together, this study prioritized a number of candidate genes, both novel hits and refining existing GWAS hits, at sCJD-associated loci aiding the identification of causal risk genes at GWAS signals by combining results from complementary eQTL and pQTL-based studies.

## Methods

### sCJD GWAS summary statistics

We used the summary statistics of the latest and the largest sCJD GWAS available from the GWAS Catalogue (GCST90001389)^8^. The discovery stage of this GWAS was performed on 17,679 samples (4,110 cases and 13,569 controls), and the summary statistics contained information on 6,314,492 high-quality imputed single-nucleotide polymorphisms (SNPs) across the autosomes^8^. As the original sCJD GWAS summary statistics were in GRCh37 human reference genome assembly and the molecular QTL catalogues and TWAS/PWAS panels used were in GRCh38 assembly, we first lifted over the variant positions from the GRCh37 to the GRCh38 genome build by using Picard (v2.22.10) LiftOver tool with “RECOVER_SWAPPED_REF_ALT=true” parameter. The SNPs that could not be lifted over to the GRCh38 genome build (7,052 SNPs; corresponding to 0.1% of total) were excluded from this study, and the remaining variants were reannotated with dbSNPv151 (GRCh38) using BCFtools annotate function. The resulting file was used in downstream molecular QTL-based analyses (e/pQTL-GWAS coloc and TWAS/PWAS) for the systematic gene prioritization pipeline.

### Gene prioritization and functional interpretation analyses

For the systematic gene prioritization pipeline we considered three domain-specific analyses, namely variant annotation, eQTL-GWAS integration, and pQTL-GWAS integration domains, for which detailed information is provided below.

### Variant annotation

We considered the index variants in each locus described in the sCJD GWAS publication^8^, namely rs3747957 in *STX6* locus, rs1799990 in *PRNP* locus, rs2267161 in *GAL3ST1* locus, rs9065 in *PDIA4* locus, and rs6498552 *BMERB1* locus for three specific criteria. First, we investigated the nearest protein-coding genes with respect to the genomic position of these lead SNPs; then we queried whether they are rare (MAF < 1% in gnomAD v4.1 non-Finnish European [NFE] samples) and/or protein-altering (missense or predicted loss-of-function) genetic variants for the nearest protein-coding genes they might reside in. Detailed information on these SNPs can be found in **Supplementary Table 1**.

### eQTL-GWAS integrative analyses

For the eQTL-GWAS integrative analyses, we processed and used publicly available bulk brain and brain cell-type-specific *cis*-eQTL catalogues and TWAS reference panels from different cohort and datasets. These included 6 bulk brain region datasets (as reanalyzed and described in detail in Bellenguez et al.^15^) of 3 AMP-AD cohorts; namely, the Mayo RNAseq Study (MayoRNAseq^19^) temporal cortex (TCX), the Religious Orders Study and Memory and Aging Project (ROSMAP^20,21^) dorsolateral prefrontal cortex (DLPFC), and The Mount Sinai Brain Bank study (MSBB^22^) Brodmann areas (BA) 10, 22, 36, and 44. Moreover, the following 4 additional bulk brain region datasets of GTEx v8 cohorts^23^ were used for eQTL-based analyses: hippocampus, frontal cortex, cortex (right cerebral frontal pole), and BA24. Furthermore, we leveraged the information cell-type-specific eQTLs (ct-eQTL) mapped in eight major brain cell types (excitatory neurons, inhibitory neurons, astrocytes, oligodendrocytes, microglia, oligodendrocyte precursor cells/committed oligodendrocyte precursors [OPCs/COPs], pericytes, and endothelial cells) from Bryois et al.^24^ and in primary microglia from Young et al.^25^ and from the Microglia Genomics Atlas (MiGA) study^26^ (medial frontal gyrus, superior temporal gyrus, subventricular zone, thalamus, and meta-analysis of four brain regions). Further information on each cohort and dataset can be found in respective publications cited and in **Supplementary Table 2**.

To investigate the potential genetic colocalization between sCJD risk association signals and eQTL/ct-eQTL signals controlling *cis* gene expression of nearby (1 Mb) genes in bulk brain and in brain cell types, we performed Bayesian colocalization analyses using coloc (v5.2.2; “coloc.abf” function with default priors)^27^ for each tested gene within above mentioned 24 distinct eQTL/ct-eQTL catalogues. The coloc analyses outputs for posterior probabilities (PPs) for five following hypotheses regarding two signals compared: H0 (no causal variant for both traits), H1 (causal variant only for sCJD GWAS), H2 (causal variant only for eQTL), H3 (two different causal variants) and H4 (common causal variant shared between sCJD GWAS and eQTL). We defined a eQTL signal as colocalized with sCJD GWAS if coloc PP4 (the posterior probability for H4) was ≥70%. Furthermore, we investigated the association between genetically regulated predicted gene expression and sCJD risk by performing TWAS in 10 bulk brain gene expression reference panels for each heritable gene expression feature. We used FUSION^28^ pipeline (using “FUSION.assoc_test.R” with default parameters) to run TWAS on 6 bulk brain custom gene expression reference panels from AMP-AD cohorts together with a custom linkage disequilibrium (LD) reference data derived from 1000 Genomes (1KG) project unrelated non-Finnish European samples (as described in detail in Bellenguez et al.^15^), meanwhile MASHR models of remaining 4 GTEx v8 brain region reference panels were used with S-PrediXcan^29,30^ (with non-default parameters “-- keep_non_rsid --model_db_snp_key varID --additional_output –throw”) implemented in MetaXcan v0.6.12 tools^29^. We determined the transcriptome-wide significance thresholds based on the Bonferroni correction on transcriptome-wide number of tested features in each gene expression reference panel (**Supplementary Table 2**). Moreover, fine-mapping of significant TWAS results was performed with Fine-mapping Of CaUsal gene Sets (FOCUS)^31^ v0.803 tool within five distinct genetic regions constructed by 1 Mb extended GWAS index variant coordinates (with “--locations” parameter), where we calculated posterior inclusion probabilities (PIPs) for TWAS associations and used these to define associations within 90% credible sets as fine-mapped TWAS associations.

### pQTL-GWAS integrative analyses

For the pQTL-GWAS integrative analyses, we accessed the publicly available bulk brain *cis*- pQTL datasets from Wingo et al.^16^ and reprocessed and reannotated these for pQTL-GWAS coloc and PWAS analyses. First, pQTL-GWAS coloc analyses were performed as described above using coloc pipeline, and by using pQTL catalogue (v2) from ROSMAP DLPFC cohort. Second, ROSMAP DLPFC (v2) and Banner Sun Health Research Institute (Banner) DLPFC PWAS reference panels were used using FUSION pipeline described above. Detailed information on these datasets and cohorts, including PWAS significance thresholds and number of samples, can be found in **Supplementary Table 2**.

### Systematic gene prioritization

To combine evidence for each candidate sCJD risk gene and nominate prioritized sCJD risk genes and related risk-associated molecular mechanisms, we applied a systematic gene prioritization and functional interpretation analysis pipeline adapted from Bellenguez et al. study^15^ for Alzheimer’s disease (AD). We first brought together all evidence for the candidate sCJD risk genes as a result of (i) variant annotation, (ii) eQTL-GWAS integration, and (iii) pQTL-GWAS integration domain analyses, each having various categories and subcategories with predetermined weighting scheme for single hits and replicated hits (across different e/pQTL coloc or TWAS/PWAS analyses), all described in detail in **Supplementary Table 3**. The weighted sum of the hits in different categories resulted in a gene prioritization score (between 0-42) for each candidate gene (i.e, a gene with a hit in at least one subcategory and with a gene prioritization score >0).

This was followed by the assignment of each candidate gene based on their genomic coordinates to 3 different types of loci and indexed: (i) the genes within 1 Mb extended coordinates of 3 genome-wide significant (GWS) index variants (with *P* ≤ 5x10^-8^) from the sCJD GWAS assigned to respective 3 GWS loci (*STX6* [G1], *PRNP* [G2], and *GAL3ST1* [G3] loci), (ii) the genes within 1 Mb extended coordinates of 2 highlighted subthreshold index variants (with *P* ≤ 5x10^-6^) from the sCJD GWAS assigned to two subthreshold loci (*PDIA4* [S1] and *BMERB1* [S2] loci), and (iii) the remaining candidate genes were grouped together if they were positioned together (<1 Mb) and these resulted in an additional 26 other loci (indexed as O1-O26). Using the pipeline described in Bellenguez et al.^15^, we then ranked all the protein-coding candidate genes in each locus based on their total weighted scores, determined the top-ranked genes, and compared the relative score differences between the top-ranked genes and the other genes in each locus to classify them as tier 1 and tier 2 prioritized risk genes, representing higher and lower levels of confidence for being true risk genes in loci, respectively. Furthermore, using a large publicly available single-nucleus RNA sequencing (snRNA-seq) study of 1.4 M nuclei from 84 human dorsolateral prefrontal cortex brain samples (The Seattle Alzheimer’s Disease Cell Atlas [SEA-AD]^32^), we first estimated average gene expression of each candidate risk gene within annotated major brain cell type clusters and then calculated the cell-type-specific gene expression proportions across 7 major brain cell types. Finally, gene set enrichment and protein-protein interaction analyses for the gene lists of tier 1 and all prioritized risk genes were performed using STRING v12^33^ with default parameters.

## Results

Our systematic gene prioritization pipeline identified 17 tier 1 prioritized risk genes and 15 tier 2 prioritized risk genes in 30 risk loci (**Fig. 1** and **Supplementary Table 4**). We could resolve all 3 GWS and 2 subthreshold sCJD risk loci with nominated tier 1 risk genes. Our integrative multiomic analyses identified candidate risk genes in another 26 loci (‘other’ loci), of which 12 harboured tier 1 prioritized risk genes.

**Figure 1.**
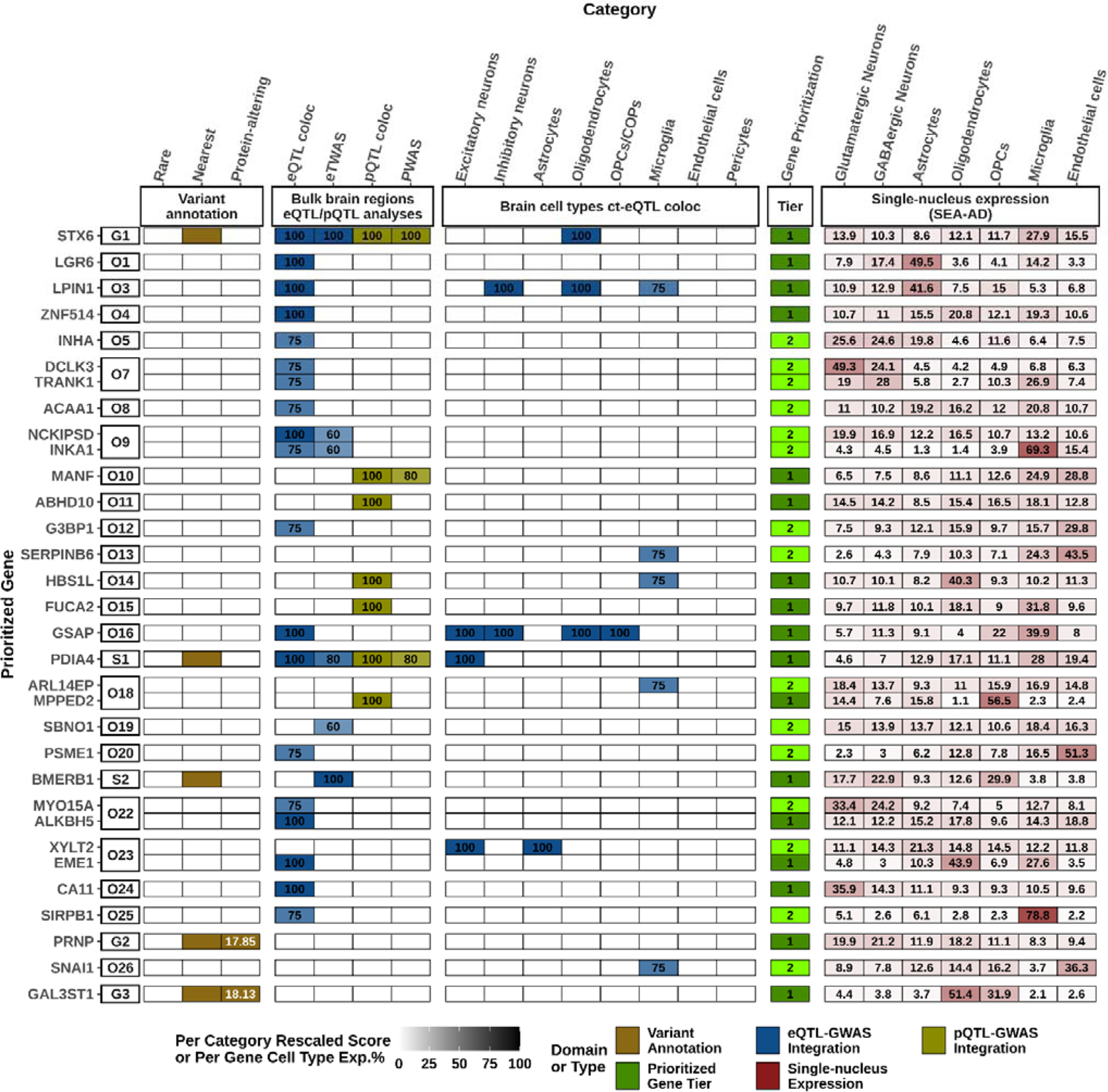
Gene prioritization results for sCJD GWAS. A visual summary of weighted evidence category scores for each prioritized risk gene, together with brain cell-type-specific gene expression proportions. The figure shows a total of 32 prioritized risk genes (17 tier 1 and 15 tier 2). The leftmost squares indicate the locus indexes where “G” is used for the genome-wide significant loci, “S” for the subthreshold loci, and “O” for the remaining other loci. The types of evidence for each category are coloured according to the three different domains to which they belonged. Weighted scores for each evidence category are rescaled to a 0–100 scale based on the maximum score a candidate gene can obtain from a category (see **Supplementary Table 3**). The darker colours represent higher scores in categories or higher average gene expression proportions in the 7 major brain cell types, while tier 1 prioritized genes are displayed in dark green and tier 2 prioritized genes are displayed in light green. Only tier 1 and tier 2 genes are shown for each locus, and all candidate genes considered and scored can be found in **Supplementary Table 4**. CADD (v1.7) PHRED scores for index variants are labelled in white within the respective squares in variant annotation domain. eQTL, expression QTL; pQTL, protein-expression QTL; ct-eQTL, cell-type-specific eQTL; coloc, colocalization; TWAS, transcriptome-wide association study; PWAS, proteome-wide association study; OPCs, oligodendrocyte precursor cells; COPs, committed oligodendrocyte precursors.

### Genome-wide significant loci

At the chromosome 1 ***STX6* locus** (G1) we observed 23 GWS SNPs. *STX6* was the nearest gene to the synonymous index variant rs3747957 and also the tier 1 prioritized gene with the highest score in this study (23), as its prioritization was supported by replicated hits in multiple subcategories (**Fig. 1-2** and **Supplementary Tables 5-9**). Remarkably, across 10 bulk brain cohorts, we found strong evidence of eQTL-GWAS colocalization (PP4s = 94.3-98.0%), in addition to having a ct-eQTL-GWAS colocalization hit specific for oligodendrocytes (PP4 = 97.7%) and a pQTL-GWAS coloc hit in DLPFC (PP4 = 99.2%) (**Fig. 2**). Moreover, the fine-mapped TWAS results showed that genetic upregulation of *STX6* was significantly associated with increased sCJD risk in multiple studies (FOCUS PIPs = 0.92-1; the most significant being in the brain region BA44; *P* = 7.92x10^-9^, Z-score = +5.77), which was also supported by protein expression level with replicated PWAS hits (*P* = 1.34x10^-8^, Z-score = +5.68 and *P* = 1.25x10^-6^, Z-score = +4.85 in the DLPFC analyses of the ROSMAP and Banner cohorts, respectively; **Fig. 3**).

**Figure 2.**
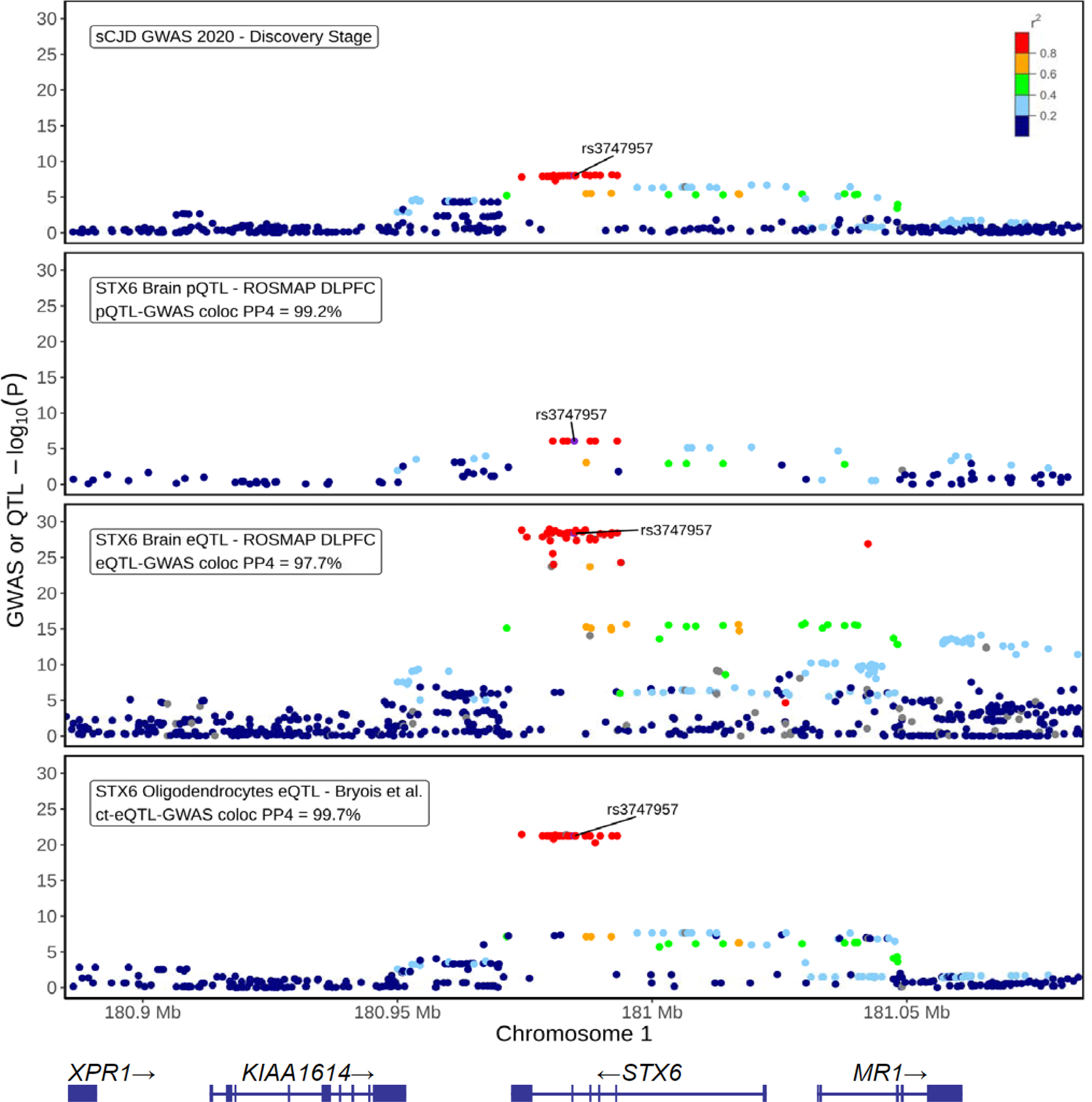
Regulation of *STX6* brain gene and protein expression by the sCJD-risk-colocalized eQTLs and pQTLs within the *STX6* locus. The regional plots of (i) sCJD GWAS association signal (*n* = 17,679), (ii) STX6 brain pQTL signal in DLPFC (ROSMAP DLPFC pQTL catalogue, *n* = 376), (iii) *STX6* bulk brain eQTL signal in DLPFC (ROSMAP DLPFC eQTL catalogue, *n* = 560), and (iv) *STX6* ct-eQTL signal in oligodendrocytes (Bryois et al.^24^ ct-eQTL catalogue, *n* = 192) are shown for 100 kb extended genomic coordinates of the *STX6* locus index variant rs3747957 (chr1:180884717-181084717). Boxes in each panel shows QTL-GWAS coloc PP4 values between the molecular QTL signal and the GWAS signal for all tested variants (see **Supplementary Tables 5-6)**. The index variant is shown in purple, and LD r^2^ values (calculated within 1 KG non-Finnish European samples [*n* = 404] with respect to the index variant) are indicated on a color scale, and variants that are not available in the LD reference panel are shown in grey. *y* axis,−log10 GWAS or QTL *P*; *x* axis, GRCh38 genomic position on chromosome 1 together with the annotation for the genomic positions of the protein-coding genes in the locus. eQTL, expression QTL; pQTL, protein-expression QTL; ct-eQTL, cell-type-specific eQTL; coloc, colocalization.

**Figure 3.**
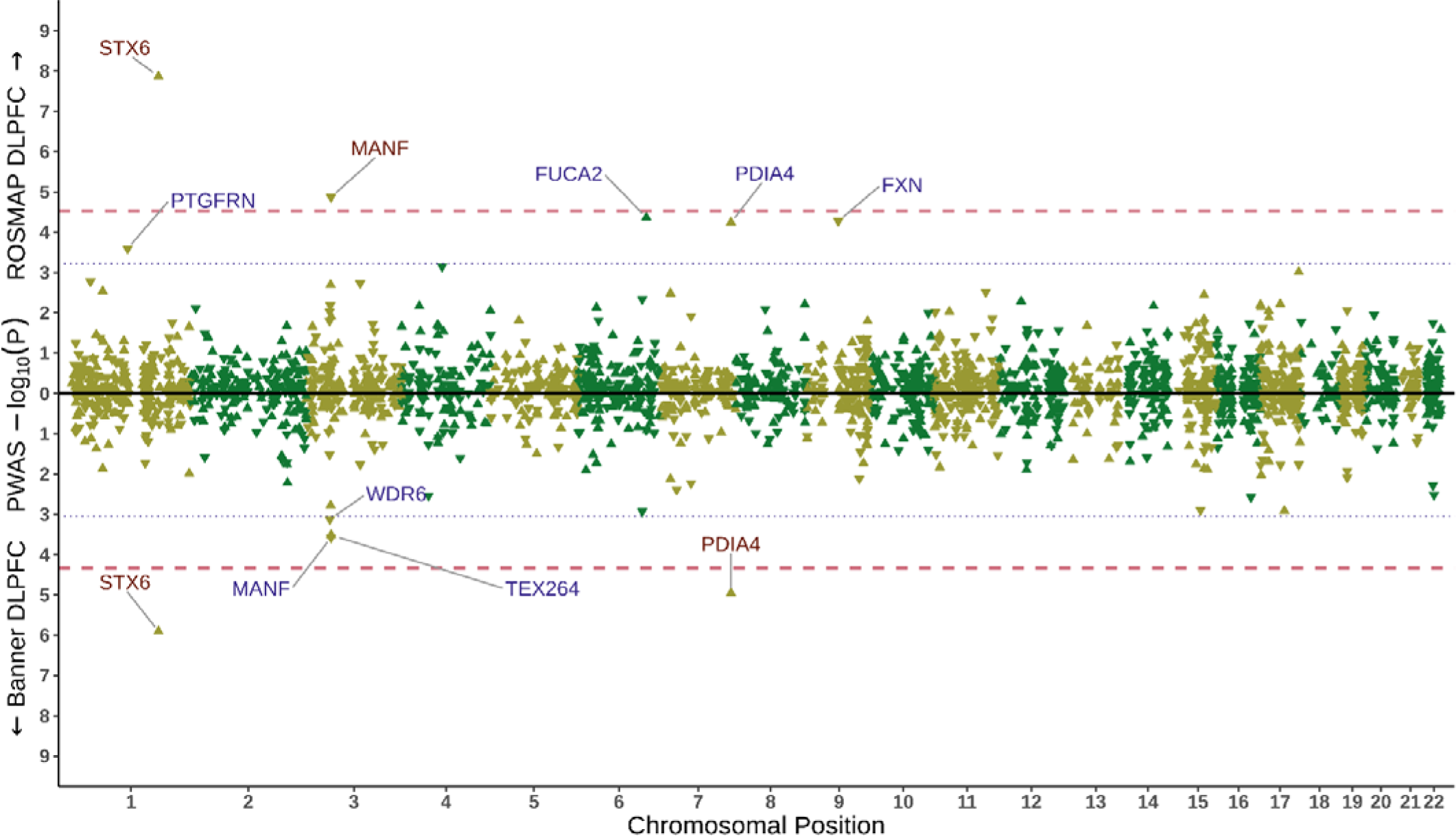
sCJD brain proteome-wide association study results. sCJD brain proteome-wide association study (PWAS) results are shown proteome-wide for both of the PWAS reference panels with two mirrored Manhattan plots on the x-axis; the upper side of the plot displays the results for ROSMAP DLPFC PWAS, while the lower side of the plot displays the results for Banner DLPFC PWAS. Proteome-wide significance thresholds (0.05 divided by number of tested associations; see **Supplementary Table 2**) for both analyses are indicated with red dashed lines and suggestive significance thresholds (1 divided by number of tested associations) with a blue dotted line, and all the genes whose protein products are passing these thresholds are labeled, and colored based on their significance (red: proteome-wide significant, blue: suggestive significant). The directionality of Z-scores of each PWAS association are represented with up-pointing triangles (positive Z-score) and down-pointing triangles (negative Z-score). *y* axis,−log10 PWAS *P*; *x* axis, GRCh38 chromosomal positions. DLPFC, dorsolateral prefrontal cortex.

At the chromosome 20 ***PRNP* locus** (G2) 16 GWS SNPs were located within *PRNP*. While we did not detect any coloc or TWAS driven evidence for any gene in this locus, we prioritized *PRNP* as the tier 1 risk gene, because the index variant rs1799990 was a common (NFE MAF = 34.3%) missense (p.Met129Val; CADD = 17.85) variant (**Fig. 1** and **Supplementary Table 1**). The same variant was also the most significant pQTL at this locus among the 183 tested. The major and protective G allele (p.129Val) was nominally associated with increased PrP levels in DLPFC (*P* = 8x10^-3^, beta = +0.019). Nevertheless, pQTL coloc results for **PRNP** showed limited pQTL-GWAS coloc (PP4 = 41%) due to the modest pQTL signal in the locus. Furthermore, no heritable PWAS models were available for PrP protein expression, thus it could not be tested in PWAS. Risk conferred by rs1799990 is complex, in that it varies between aetiological types of prion disease^34–37^, but the heterozygous genotype is strongly associated with reduced risk of sCJD and more slowly progressive phenotypes relative to both homozygous genotypes^34^. The biological relevance of this pQTL is therefore unclear.

At the chromosome 22 ***GAL3ST1* locus** (G3) we found 2 GWS SNPs centering *GAL3ST1* as the nearest gene. *GAL3ST1* could be prioritized as the tier 1 risk gene, as the index variant rs2267161 (p.Met29Val, CADD score 18.13) was a common (NFE MAF = 31.1%) missense variant, despite the presence of two other candidate genes in the locus: *TCN2* (50 kb downstream from the index variant with fine-mapped TWAS hit in BA22) and *INPP5J* (>500 kb downstream from the index variant with a borderline eQTL coloc hit in DLPFC in the ROSMAP cohort) (**Fig. 1** and **Supplementary Tables 5 and 7**). Moreover, *GAL3ST1* p.Met29Val index variant has strong associations with sulfatide (SHexCer) blood lipids (five different classes and total SHexCer, *P*=2.5x10^-15^ – 2.7x10^-37^) with the sCJD risk allele rs2267161-C conferring increased lipid levels ^38^.

### Subthreshold loci

At the chromosome 7 ***PDIA4* locus** (S1), which was a hit in gene-wide analyses in the previous study^8^, the GWAS association signal surrounded *PDIA4* with a minimum *P* of 1.66x10^-6^ for the 3’ UTR index variant rs9065. We detected multiple lines of evidence supporting *PDIA4* (gene prioritization score of 21, the second highest in this study after *STX6*) as a tier 1 prioritized in this locus (**Fig. 1** and **Supplementary Tables 5-9**). Across 8 bulk brain cohorts, we found strong evidence of eQTL and sCJD risk colocalization (PP4s between 85.1%-96.1%). This appeared to be largely driven by excitatory neurons (PP4 = 76.2%). *PDIA4* was also a fine-mapped TWAS hit (FOCUS PIP = 98.9%, *P* = 1.02x10^-6^, Z-score = +4.89) and a significant PWAS hit in Banner DLPFC (*P* = 1.1x10^-5^, Z-score = +4.39; **Fig. 3**). Genetic upregulation of both transcript and protein expression confer increased risk of sCJD. Finally, *PDIA4* pQTLs also colocalized with the sCJD GWAS (PP4 = 94.7%).

At the chromosome 16 ***BMERB1* locus** (S2), the intronic index variant rs6498552 was close to the GWS threshold (rs6498552 *P* = 5.73x10^-8^) ^8^. *BMERB1* (formerly known as *C16orf45*) was the only candidate gene in S2 and we prioritized it as a tier 1 risk gene, as it had replicated fine-mapped TWAS hits in GTEx Frontal Cortex (*P* = 4.7x10^-6^, Z-score = -4.58, FOCUS PIP = 96.5%) and GTEx Hippocampus (*P* = 4.7x10^-6^, Z-score = -4.58, FOCUS PIP = 96.9%) analyses where the predicted gene expression was conversely associated with the risk of sCJD (**Fig. 1** and **Supplementary Table 7**).

### Other loci

Of the remaining 26 ‘other’ loci, 22 had protein-coding genes in which we performed gene prioritization analysis. Of note, variant annotation domain does not contribute to gene prioritization in these loci because they do not harbour GWAS index variants^8^. Nevertheless, we could assign a tier 1 prioritized risk gene in 12 of these 22 loci. Moreover, for the remaining 10 risk loci, 8 had a single tier 2 prioritized risk gene and 2 (O7 and O9) had two tier 2 prioritized risk genes with similar weighted gene prioritization scores (**Fig. 1** and **Supplementary Table 4**). While full results on these prioritized genes are available in **Supplementary Table 4**, below we highlight 5 of these loci containing the five highest scoring candidate genes (gene prioritization scores ≥7; all supported by hits in multiple subcategories, see **Fig. 1**), in addition to *SIRPB1* in O25 with considerable GWAS evidence.

In locus O10 we identified *MANF* as tier 1 prioritized risk gene, which was also the highest scoring gene (gene prioritization score of 8) among the other loci candidate genes. *MANF* had a pQTL-GWAS coloc hit (PP4 = 88.1%) and PWAS hit (*P* = 1.35x10^-6^, Z-score = -4.35; **Fig. 3**) in DLPFC in the ROSMAP cohort, where genetic downregulation of protein expression was associated with increased risk of sCJD. Moreover, *LPIN1* (O1) and *GSAP* (O16) tier 1 prioritized risk genes both notably exhibited replicated bulk brain eQTL-GWAS coloc hits (in 9 and 7, across 10 analyses, respectively) and also had ct-eQTL-GWAS coloc hits in multiple brain cell types (3 and 4, across 8 cell types), showing the importance of sCJD risk-associated genetic variation in both loci in terms of gene expression regulation across multiple brain regions and cell types. In locus O14, *HBS1L* was identified as the tier 1 prioritized gene through a pQTL-GWAS coloc hit in DLPFC (PP4 = 86.1%) and borderline microglia ct-eQTL-GWAS coloc hit in meta-analysis of the MiGA data (PP4 = 70.2%). Furthermore, locus O9 had 4 protein-coding candidate genes (the highest among all loci), in which two genes were prioritized as tier 2 risk genes as the weighted evidence was similar: *NCKIPSD* and *INKA1* (formerly known as *FAM212A*), positioned furthest away from each other in the locus (>1.1 Mb), had both eQTL-GWAS coloc and TWAS hits. *NCKIPSD* scored one point higher than *INKA1* because of having replicated eQTL-GWAS coloc hits (9 out of 10 analyses), although coloc PP4 for *INKA1* in DLPFC in the ROSMAP cohort was higher (98.8% vs 87.8%). Finally, in locus O25, located >3 Mb upstream of *PRNP*, *SIRPB1* was prioritized as a tier 2 risk gene as a result of an eQTL-GWAS coloc hit in BA10 (PP4 = 86.1%). Of note, *SIRPB1* had the second most significant GWAS *P* evidence among other loci candidate genes after the genes within locus O9, as the GWAS *P* for its 3’UTR variant rs2422615 was 5.26x10^-6^ (**Fig. 1** and **Supplementary Tables 5-9**).

### Gene set enrichment and interaction analyses

Using STRING, we performed gene set enrichment and protein-protein interaction analyses for the gene lists of tier 1 and all prioritized risk genes. While no significant pathways (FDR < 0.05) were found to be enriched when corrected for multiple comparisons, we detected a strong protein-protein interaction relationship between *PDIA4* and *MANF* on the basis of experimental/biochemical data, co-expression, and mentions of both genes in abstracts in the literature (see Discussion). There was also some suggestive evidence for protein-protein interactions between *PRNP* and *SIRPB1, TRANK1* and *DCLK3,* as well as *LPIN1* and *ACAA1*, although none of these were based on human experimental/biochemical data.

## Discussion

Transcriptome and proteome-wide association studies (TWAS and PWAS) and molecular QTL-GWAS colocalization analyses can contribute to a better understanding of genetic risks for diseases through refining hypotheses about implicated genes, direction of effects, cell types and pathways using GWS and subthreshold findings. Human prion diseases have not previously been studied in this way and, beyond the prion protein locus itself, suffer from a paucity of genetically validated targets for therapeutic development. In 2020, a large GWAS study in the prion disease field led to the discovery of three proposed genetic loci associated with sCJD risk^8^ in or near to *PRNP, STX6* and *GAL3ST1*, and we highlighted two subthreshold loci (*PDIA4* and *BMERB1*). We aimed to harness transcriptomic and proteomic datasets to provide further insight into sCJD risk. Herein we report considerable molecular QTL-based evidence that supports a causal role for genetically upregulated syntaxin-6 gene and protein expression in risk of sCJD relative to other genes at the locus, and a cell-type-specific relevance of the GWAS signal in regulating *STX6* gene expression in oligodendrocytes but not in other brain cell types. Furthermore, both subthreshold hits we previously highlighted, *PDIA4* and *BMERB1*, also show significant associations between their genetically regulated expression and sCJD risk. We also found that reduced protein expression of a further gene product, previously unconnected to prion diseases, *MANF*, was associated with increased sCJD risk in PWAS. Interestingly, the sCJD proposed risk gene *GAL3ST1* encodes an enzyme involved in the synthesis of sulfatides, which are a major lipid component of the myelin sheath and are known to have experimental links with both *MANF* and ER stress^39^, providing an indirect link to *PDIA4*. This work therefore refines and proposes new hypotheses about mechanisms of risk in human prion diseases.

Variants in and near to the syntaxin-6 (*STX6*) gene are genetic risk factors for sCJD^8^ and the most common primary tauopathy, progressive supranuclear palsy (PSP)^40–44^. Syntaxin-6 is a member of the SNARE protein family^10^, which mediate the final step of membrane fusion during vesicle transport, and thus its identification in GWAS implicated intracellular trafficking as a causal disease mechanism. However, although *STX6* appears to modify disease susceptibility^8^, in more recent work we have shown there is no association with age of onset or disease progression^45^, and knockout of *Stx6* expression in mouse has no, or modest effects, on prion disease incubation time^46^. In this work, we show increased *STX6* expression was significantly linked to risk of sCJD across multiple reference panels both for TWAS and PWAS, along with e/pQTL-GWAS colocalization, whereas evidence was limited for other genes (including *KIAA1614*) at the same locus. These findings are concordant with previous studies in tauopathies correlating genetic risk loci with transcriptomic and proteomic data. Indeed, using reference data from the GTEx Consortium, a PSP TWAS study identified that the *STX6* risk haplotype was associated with differential expression of the gene^41^. Furthermore, a recent frontal cortex case–control EWAS meta-analysis identified *STX6* as being hypomethylated at CpG sites in PSP compared to controls^47^. Interestingly, *STX6* has also been identified as conferring Alzheimer’s disease (AD) risk in a recent AD PWAS study, with increased syntaxin-6 protein levels in the brain being causally associated with the disease^48^. We conclude that syntaxin-6 has pleiotropic risk effects in neurodegenerative diseases, which are driven by a common genetic mechanism of increased protein expression.

As expected, *PRNP* and *GAL3ST1* were not identified as PWAS or TWAS hits, which is in keeping with the candidate mechanisms of these genes being driven by common missense variants. At *PRNP*, the p.Met129Val polymorphism is known to be a strong modifier of prion disease determining predisposition to sCJD^49^ and iatrogenic CJD (iCJD) ^50^, as well as influencing age of disease onset and/or disease progression in kuru^37^ and some inherited prion diseases^36^, where in general the heterozygous genotype is protective compared to both homozygous genotypes. It is important to note that most molecular QTL studies, including the ones used in our study, are based on additive models (where the effect of increasing number of alleles are tested against the molecular phenotype outcome), therefore this can be one of the limiting factors for finding significant downstream effects of this genetic variant on PrP expression. Moreover, codon 129 has complex effects, exemplified by susceptibility to variant CJD (vCJD), the human form of bovine spongiform encephalopathy, with all but one definite case being homozygous for methionine at codon 129^51^. These human associations correlate well with modelling of the codon 129 genotype in mouse^52^ and are in keeping with a mechanism of codon 129 genotypic risk that involves the selection of prion strains and dominant negative effects. Galactose-3-O-sulfotransferase 1 (*GAL3ST1)* is an oligodendrocyte expressed enzyme, which catalyses the sulfation of Golgi-membrane sphingolipids to form sulfatides. These are important lipids in the brain and essential constituents of the myelin sheath^11^. In the *GAL3ST1* gene, a common amino acid variant (p.Val29Met) confers increased risk of sCJD. In recent lipidomics GWAS studies the p.Val29Mel variant was associated with altered concentrations of blood sulfatides ^38,53^. Therefore, as there is already strong evidence for a genetic mechanism at both of these loci independent from expression change, we would not expect either *PRNP* nor *GAL3ST1* to be a TWAS/PWAS hit. Of note, *TCN2*, upstream at the *GAL3ST1* locus, was identified as a fine-mapped TWAS hit in a single cohort, and is therefore an alternative albeit lower priority candidate at the locus.

Previously, we reported suggestive evidence that the *PDIA4* locus was associated with sCJD risk by gene-based testing in the discovery stage of GWAS^8^. These TWAS and PWAS analyses provide an additional, complementary approach to explore the association of the *PDIA4* locus with sCJD risk. *PDIA4* was both TWAS and PWAS significant with a consistent positive Z-score suggesting genetic upregulation of this gene increases risk for sCJD, supported also by the replicated e/pQTL-GWAS colocalization. *PDIA4* encodes a member of the protein disulphide isomerase (PDI) family of proteins and is localised to the endoplasmic reticulum (ER) where it mediates oxygen-dependent disulphide bond formation and consequently the correct folding of both transmembrane and secreted proteins^54^. It has broad brain expression and its function has been linked to the unfolded protein response (UPR). Interestingly, PDIA4 has been implicated in prion disease pathogenesis^55^ as well as independently emerging as a central, generic player in other neurodegenerative diseases (reviewed in^56^) suggesting it may have risk effects across multiple protein misfolding diseases. Specifically, the PDI gene family is upregulated in prion-infected cultured cells as well as in prion-infected hamster brains early in disease pathogenesis, which progressively increases at later stages of the disease^55^. This is further supported by two further independent studies showing *Pdia4* is upregulated both at the RNA and protein level in mice infected with RML prions^57^.

The identification of *PDIA4* as a TWAS/PWAS hit localising to excitatory neurons (through the ct-eQTL-GWAS coloc analyses) further implicates the UPR in human sCJD. Although the UPR is a physiologically protective cellular response, which protects against ER stress driven by the accumulation of misfolded proteins or other stressors^58^, dysregulation of the UPR across multiple neurodegenerative diseases leads to translational failure ultimately culminating in neuronal loss^59–61^. This translational failure is driven by the phosphorylation of the α-subunit of eukaryotic translation initiation factor, eIF2α^62^. Importantly, the UPR has been highlighted as a mechanism in prion disease pathogenesis, with eIF2α-P driving persistent translational repression of global protein synthesis in prion-infected mice, leading to synaptic failure and neuronal loss^63^. In a more recent study it has been shown that the protracted UPR typical of prion diseases also induces diacylation of a key phosphoinositide kinase, PIKfyve, resulting in its degradation and consequently endolysosomal hypertrophy and activation of TFEB-dependent lysosomal enzymes^64^. This has been proposed to underpin a defining histopathological trait of sCJD: spongiform degeneration. Therefore, the identification of *PDIA4* in this study, and its strong links to the UPR, are in keeping with the emerging theme in the prion disease field that a dysregulated UPR is a driver of neurotoxicity.

Continuing with this theme, Mesencephalic Astrocyte-derived Neurotrophic Factor (*MANF*), also implicated in the ER stress response, was a PWAS and pQTL-GWAS coloc hit. Although it did not surpass the stringent threshold of significance in the Banner DLPFC PWAS reference panel (*P* = 3x10^-4^, Z-score = -3.65; **Fig. 3**), this analysis supported the same direction of effect at a suggestive significance level and its conserved position in the top three most significant hits across panels provides confidence its levels are associated with risk of the disease. Mammalian *MANF* was first reported to have neurotropic effects on dopaminergic neurons^65^, promoting their survival^66^. It has particularly high expression in the brain (reviewed in^67^) with ER stress promoting its upregulation^68^ as well as its secretion into the extracellular environment^69,70^. *MANF* has been shown to be an important regulator of the UPR^68,71^, which is further supported with studies using mice with *Manf* knockout which show abnormal activation of the UPR^72^. Interestingly, it has recently been shown that human MANFs directly bind to sulfatide promoting the cellular uptake of MANF, which alleviates the ER stress response in cells thereby conferring cytoprotection^39^. Its identification in this study as a new candidate gene therefore provides potential convergence with another sCJD risk gene, *GAL3ST1*. Additionally, as a secreted factor from astrocytes, it provides support for the increasingly accepted notion that the interplay between astrocytes and neurons in prion disease is a key pathogenic phenomenon^73^.

Interestingly *SIRPB1*, located >3 Mb upstream of *PRNP* and genetically linked to *PRNP*, was prioritized as a tier 2 risk gene with there being suggestive evidence for a protein-protein interaction between *PRNP* and *SIRPB1*. *SIRPA* encodes signal regulatory protein α (SIRPα), a protein enriched in microglia which plays a key modulatory role of phagocytosis. However, SIRPα does not appear to play a role in prion pathogenesis *in vivo*^74^.

Another fascinating finding that came out of this study comes from analysis of cell-type-specific eQTLs (ct-eQTLs), which revealed striking cell-type-specific effects in the genetic control of *STX6* gene expression by risk variants with the *STX6* signal specifically colocalizing with oligodendrocyte eQTLs (**Fig. 2**). This provides suggestive evidence that *STX6* may be exerting its risk effects in oligodendrocytes. Oligodendrocytes are an understudied cell population in the prion disease field, but one study provided evidence that oligodendrocytes do not replicate prions and are resistant to prion infection^75^. However, it is possible that the relationship between neurons, oligodendrocytes and other brain cell types is crucial for prion formation, propagation, clearance or neurotoxicity. Indeed, there is suggestive evidence for a role of oligodendrocytes in prion disease through dysregulation of oligodendrocyte-specific genes in transcriptomic studies^76–78^. Furthermore, a recent study showed that NG2 glia, oligodendrocyte-lineage cells, exert a protective effect against prion-induced neurotoxicity by interacting with microglia and inhibiting critical signalling pathways^79^. It is also noteworthy that in human patients, oligodendroglial PrP pathology has been reported in certain histotypes of sCJD^80^. Therefore, oligodendrocytes may be implicated in prion pathogenesis, which is further supported by the convergence of the two non-*PRNP* sCJD risk factors, *STX6* and *GAL3ST1*, in this cell type.

This study has also several limitations. Firstly, our molecular QTL-based analyses were limited to eQTLs and pQTLs; however, the inclusion of other molecular QTLs such as splicing QTLs (sQTLs), methylation QTLs (mQTLs), and histone acetylation QTLs (haQTLs) in future studies could provide additional sCJD risk-associated molecular mechanisms, which can be complementary in terms linking the GWAS signals to similar sets of prioritized risk genes or to other candidates. Secondly, the molecular QTL-based analyses we used were designed to capture GWAS-relevant regulatory variants for the features in *cis* (typically within a window of < 1 Mb from the features), yet GWAS signals could be related to *trans*-QTLs, linking associations to distant candidate genes. However, generation of *trans*-eQTL and *trans*-pQTL catalogues have been historically difficult due to multiple problems related to sample size and control of confounders^81^, although there has been recent progress in large-scale brain trans-eQTL catalogues ^82^, opening up new analysis opportunities in the future for rare cases where a GWAS signal is acting through a *trans*-eQTL signal. Thirdly, despite the recent progress in availability of brain ct-eQTL catalogues^24,83^, no such cell-type-specific pQTL catalogues are available to our knowledge; but the latest advances in the field for single-nucleus proteomics^84^ may lead to brain ct-pQTL datasets in the foreseeable future.

In conclusion, our results are compatible with the leading hypotheses for the three known genetic risk factors for sCJD, with there being robust evidence for increases in *STX6* expression driving disease risk, but not for *PRNP* and *GAL3ST1*, which are thought to be driven by missense SNPs. Furthermore, this functionally-informed analysis of sCJD GWAS summary statistics provides additional suggestive evidence and connections between other prioritized genes, including *PDIA4, BMERB1* and *MANF,* and generally, for a role of glial cells and the UPR in sCJD aetiology (**Fig. 4**). Future functional studies may confirm the target prioritized sCJD risk genes and risk-associated molecular mechanisms highlighted in our study, leading to better understanding of the disease mechanisms and consequently providing new therapeutic opportunities for sCJD, with potential relevance to other neurodegenerative diseases.

**Figure 4.**
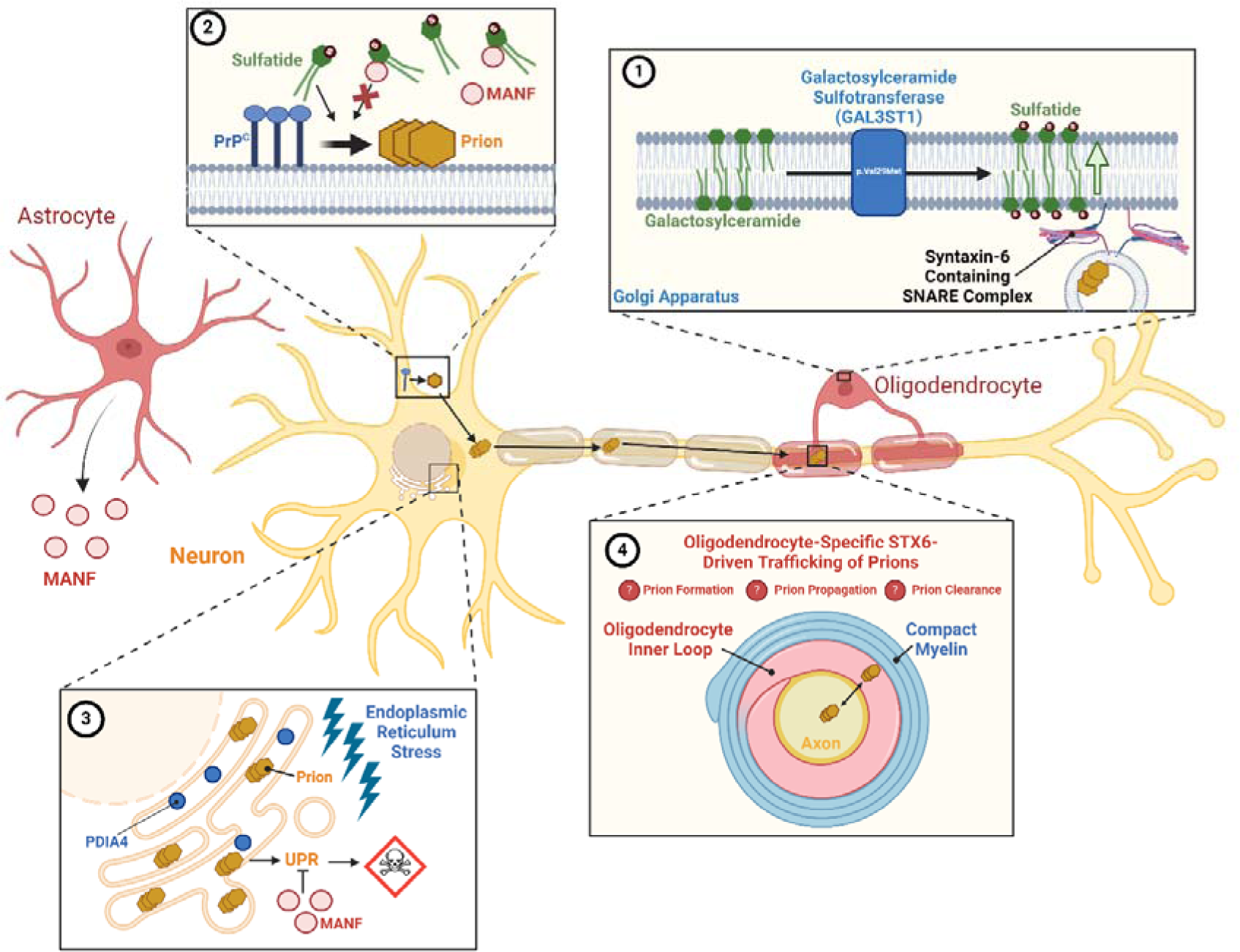
Speculative Model of the Cell Types and the Potential Relationship Between Prioritized Risk Genes and their Mechanisms. A common amino acid variant (p.Val29Met) in the *GAL3ST1* gene, encoding galactosylceramide sulfotransferase, increases sulfatide production predominantly in oligodendrocytes, conferring increased risk of sCJD (**1**). Sulfatide may act as a cofactor in PrP^C^ conversion or prion propagation, which may be intercepted by the astrocyte-secreted factor, MANF, which binds to sulfatide extracellularly (**2**). Sulfatide may additionally promote the cellular uptake of MANF allowing it to work in concert with PDIA4 to protect against the adverse effects of ER stress and the sustained unfolded protein response characteristic of prion infection (**3**). Increased syntaxin-6 expression predominantly in oligodendrocytes may be altering the trafficking of either PrP^C^ or prions with implications on prion formation, propagation and/or clearance (**4**). Figure created on Biorender.

## Data availability and URLs

The sCJD GWAS^8^ summary statistics is available at the European Bioinformatics Institute GWAS Catalog portal (https://www.ebi.ac.uk/gwas/) under accession no. GCST90001389.

SEA-AD^32^ brain single nucleus gene expression matrices (https://registry.opendata.aws/allen-sea-ad-atlas/)

Full e/pQTL-GWAS coloc and TWAS/PWAS results from this study are available at https://doi.org/10.5281/zenodo.12507355, while significant-only results are shown in **Supplementary Tables 5-9**.

Molecular eQTL and pQTL related datasets used in this study are publicly available (see also **Supplementary Table 2**):

eQTLs and TWAS reference panels in AD-relevant bulk brain regions from AMP-AD cohorts, as analyzed by Bellenguez et al.^15^: (https://doi.org/10.5281/zenodo.5745927);

GTEx v8^23^ eQTL catalogues (https://www.gtexportal.org/);

GTEx v8 MASHR^29,30^ expression prediction models for TWAS (https://predictdb.org/post/2021/07/21/gtex-v8-models-on-eqtl-and-sqtl/#mashr-based-models);

Bryois et al.^24^ ct-eQTL catalogues (https://doi.org/10.5281/zenodo.5543734);

MiGA eQTL catalogues (https://doi.org/10.5281/zenodo.4118605 and https://doi.org/10.5281/zenodo.4118676);

Wingo et al.^16^ v2 pQTL catalogues & PWAS reference panels (https://www.synapse.org/#!Synapse:syn23627957).

## Funding and Acknowledgements

The work was funded by the Medical Research Council (UK). SM and JC are National Institute for Health Research (NIHR) Senior Investigators (JC is emeritus). FK receives a postdoctoral fellowship (BOF 49758) from the University of Antwerp Research Fund.

The data available in the AD Knowledge Portal would not be possible without the participation of research volunteers and the contribution of data by collaborating researchers. The results published here are in whole or in part based on data obtained from the AD Knowledge Portal (https://adknowledgeportal.org). Data generation was supported by the following NIH grants: P30AG10161, P30AG72975, R01AG15819, R01AG17917, R01AG036836, U01AG46152, U01AG61356, U01AG046139, P50 AG016574, R01 AG032990, U01AG046139, R01AG018023, U01AG006576, U01AG006786, R01AG025711, R01AG017216, R01AG003949, R01NS080820, U24NS072026, P30AG19610, U01AG046170, RF1AG057440, and U24AG061340, and the Cure PSP, Mayo and Michael J Fox foundations, Arizona Department of Health Services and the Arizona Biomedical Research Commission. We thank the participants of the Religious Order Study and Memory and Aging projects for the generous donation, the Sun Health Research Institute Brain and Body Donation Program, the Mayo Clinic Brain Bank, and the Mount Sinai/JJ Peters VA Medical Center NIH Brain and Tissue Repository. Data and analysis contributing investigators include Nilüfer Ertekin-Taner, Steven Younkin (Mayo Clinic, Jacksonville, FL), Todd Golde (University of Florida), Nathan Price (Institute for Systems Biology), David Bennett, Christopher Gaiteri (Rush University), Philip De Jager (Columbia University), Bin Zhang, Eric Schadt, Michelle Ehrlich, Vahram Haroutunian, Sam Gandy (Icahn School of Medicine at Mount Sinai), Koichi Iijima (National Center for Geriatrics and Gerontology, Japan), Scott Noggle (New York Stem Cell Foundation), Lara Mangravite (Sage Bionetworks). Study data were generated from postmortem brain tissue obtained from the University of Washington BioRepository and Integrated Neuropathology (BRaIN) laboratory and Precision Neuropathology Core, which is supported by the NIH grants for the UW Alzheimer’s Disease Research Center (P50AG005136 and P30AG066509) and the Adult Changes in Thought Study (U01AG006781 and U19AG066567). This study is supported by NIA grant U19AG060909.

